# AI Adoption for NCDs in Kenya: A Qualitative Study

**DOI:** 10.64898/2026.05.26.26354008

**Authors:** Jessica Rayo, Wanjiru Cushny, Martin Mwangi, Steven Wanyee, Marius George Linguraru, Nelly Nyaga, Hillary Koros, Miriam Bosire, Melvine Obuya, Christine Ngaruiya

**Author notes:** **Correspondence to: Jessica Rayo,**.

## Abstract

**Background:** Non-communicable diseases (NCDs) represent a critical public health challenge in Kenya, responsible for over 50% of inpatient admissions and 40% of deaths. While digital health tools and artificial intelligence offer promising ways to improve prevention, diagnosis, and management, little is known about how these tools are perceived and used in practice. There is limited research exploring the views and lived experiences of young people in Kenya, who are a strategic priority for NCD prevention because behavioral risk factors are established in this window, and for Community Health Providers (CHPs) who provide health services within the community. This study aims to address this gap by examining the perspectives of the burden of non-communicable diseases and the potential role of digital health technologies, including artificial intelligence, for preventing and managing these conditions in these specific populations.

**Methods:** A qualitative research design using focus group discussions (FGDs) was employed in Nairobi (urban) and Busia (rural) counties between March and July 2024. Eight FGDs were conducted with 60 participants purposively sampled from three stakeholder groups: community health promoters (CHPs), healthcare workers (HCWs), and youth aged 18-35 years. A semi-structured guide, co-developed with a Community Advisory Board, explored beliefs about NCDs, health-seeking behaviors, lifestyle practices, and attitudes toward digital health and AI. Audio recordings were transcribed verbatim, translated where necessary, and analyzed thematically using grounded theory principles on NVivo software (v12).

**Results:** Six consolidated themes emerged: (1) understanding of NCDs and perceived risk; (2) barriers to NCD prevention and care; (3) the role of CHPs; (4) adoption of AI tools for NCD management; (5) trust, ethics and access concerns; and (6) community-driven recommendations for AI integration. Significant barriers including stigma, economic constraints, and barriers to care were documented alongside enthusiasm for AI tools among youth and CHPs in both urban and rural areas.

**Conclusion:** This study shows that AI tools are being used for NCD prevention and management through spontaneous community adoption. However, it emphasizes the need for culturally relevant, equitable, and community-driven solutions. Effective scaling requires the identification and bridging of digital literacy gaps, the establishment of affordable infrastructure, the protection of data privacy, and the integration of artificial intelligence tools into existing community health frameworks. This process should involve the collaboration of trusted intermediaries, such as CHPs and community leaders, to ensure successful outcomes. Future initiatives should prioritize participatory design, policy frameworks for ethical governance, and targeted capacity building to enhance acceptance and sustainability of digital health innovations in low- and middle-income country settings.

## Introduction

### The Global and Regional Burden of NCDs

Non-communicable diseases (NCDs) such as diabetes, heart disease, stroke, chronic lung disease, and cancer account for 74% of all global deaths, with 73% of NCD-related mortality occurring in low- and middle-income countries (LMICs) (1,2). In Kenya, NCDs are responsible for over 50% of inpatient admissions and 40% of hospital deaths, with annual healthcare expenditures projected to reach KES 607 billion (∼US$5.5 billion) by 2030, as new cases of major NCDs, such as hypertension, more than double from 5.3 million to over 10 million adults by 2035 (3,4). Despite the magnitude of the problem, an estimated 80% of NCD cases are preventable (1,2). This underscores the pressing need for scalable healthcare solutions to support the management of chronic conditions in LMICs and beyond (5, 2).

### Young People as Both Targets and Agents of Change

Youth represent a critical demographic for understanding and addressing noncommunicable diseases. Targeting young people is essential because behavioral risk factors responsible for at least two thirds of premature deaths due to NCDs, including tobacco use, physical inactivity, unhealthy diet, and harmful alcohol consumption, are often established during adolescence and young adulthood (6, 7). Hence, this population sits at a crucial intervention point where health trajectories can still be modified (7). Digital media and AI tools offer opportunities to reach young people through platforms they already use, delivering personalized health information in formats that resonate with their communication preferences. Understanding how youth currently engage with these technologies for NCD-related information, and identifying gaps in their knowledge and practices, is fundamental to developing effective digital health interventions that can help them modify their risk before unhealthy behaviors become entrenched.

Beyond their role as beneficiaries of prevention efforts, young people are uniquely positioned as agents of change in the fight against NCDs (8). This is because today’s youth possess unprecedented access to information through digital platforms and the capacity to mobilize communities, influence peers, and hold policymakers accountable. As highlighted in the 2011 Political Declaration on NCDs, young people can advocate for policies that improve NCD prevention and care, contribute to public education initiatives, and provide novel perspectives on prevention strategies through their command of new media channels (8).

By examining young people’s knowledge, attitudes, and practices regarding AI for NCDs, this study provides essential insights into how to harness their potential as advocates, educators, and innovators in NCD prevention. This transforms them from a target population into active partners in creating the systemic changes needed to reduce the global burden of noncommunicable diseases.

### Emergence and Promise of Digital Health and AI in LMICs

The rapid advancement of artificial intelligence (AI) in medicine is set to reshape healthcare delivery, offering both significant opportunities and complex challenges. As these technologies become more integrated into clinical practice, it is essential to address critical concerns related to patient safety, resource distribution, and health equity. In Africa, the expansion of digital health is closely tied to broader trends, including increased internet connectivity, widespread mobile phone adoption, and improvements in adult literacy (9). Research shows that regions with higher technological development tend to experience lower burdens of disease, highlighting the potential of digital health and AI to reduce health disparities and improve access for underserved populations (7). Consequently, leveraging digital technology and AI emerges as one of the most promising strategies for improving equity and access to healthcare for marginalized communities.

The Kenyan context makes this opportunity particularly salient. Kenya ranks among the highest per-capita users of ChatGPT globally reflecting a population that is not only digitally connected but actively engaging with AI tools for information-seeking, including, increasingly, health-related queries (10). This pattern of AI adoption among Kenyan youth provides both the rationale and the urgency for this study.

Digital innovations are particularly well-suited to address persistent inequities in healthcare, including limited access to reliable health information and specialized medical expertise. Generative AI, primarily through large language models (LLMs), holds promise for overcoming these barriers in LMICs by enabling scalable, context-sensitive solutions that can support both patients and providers. To fully realize these benefits, however, implementation strategies must prioritize equity, cultural relevance, and robust governance to ensure that technological progress translates into meaningful improvements in health outcomes for marginalized communities.

## Methods

### Study Design

This study employed a qualitative research design using focus group discussions (FGDs) to explore perceptions and lived experiences related to NCDs and the potential role of AI in health promotion. This approach enabled rich, context-specific insights that reflect community perspectives and realities.

### Study Setting and Participants

The study was conducted in Nairobi (urban) and Busia (rural) counties in Kenya to capture diverse socioeconomic, demographic, and geographic perspectives. Participants were purposively selected from three stakeholder groups: community health promoters (CHPs), healthcare workers (HCWs), and youth aged 18–35 years. These groups were selected for their unique and complementary roles in health education, service delivery, and digital engagement.

### Sampling and Recruitment

Purposive sampling was used to ensure diversity across gender, geography, and health system roles. Recruitment was facilitated through community networks, local leaders, health facilities, and academic institutions. A total of eight FGDs were conducted (four in each county) with 6 to 10 participants per session, totaling 60 participants.

### Data Collection

FGDs were held between March and July 2024 using a semi-structured guide co-developed by the research team and a Community Advisory Board (CAB). The CAB consisted of local stakeholders, including CHPs and youth representatives, who offered insights to align the discussion guide with community norms and priorities. The CAB provided input on key discussion topics and helped ensure contextual and cultural relevance. Topics included knowledge and beliefs about NCDs, health-seeking behavior, lifestyle practices, perceptions of AI in health, and related ethical concerns. Sessions were conducted in English or Swahili based on participant preference and facilitated by trained qualitative researchers. Each session lasted 60–90 minutes and was audio-recorded with participants’ informed consent.

### Data Analysis

Audio recordings were transcribed verbatim and translated into English where necessary. Thematic analysis was conducted using NVivo (version 12) software. Themes were developed based on recurring patterns and their relevance to the study objectives. Coding and theme development followed an iterative process, supported by team-based discussions and peer reviews, to enhance consistency and minimize bias. Final codes were then refined into broader categories and synthesized into key themes, reflecting the experiences and perspectives of participants.

### Ethical Considerations

The study received ethical approval from the AMREF Ethics and Scientific Review Committee (Ref: ESRC P1518/2023), NACOSTI (NACOSTI/P/24/31792), and County Health Offices in Nairobi and Busia. All participants provided written informed consent. Transcripts were anonymized, and data were stored on encrypted, access-restricted servers. Participants received reimbursement for time and transportation.

## Results

Five main themes emerged from the focus group discussions, revealing how Kenyan communities understand NCDs and perceive AI’s potential role in health promotion.

**Table 1:** A table showing the themes, subthemes and illustrative quotes

| Theme | Sub-theme | Illustrative quotes |
| --- | --- | --- |
| 1. Cultural and biomedical understanding of | Awareness of NCD types (diabetes, hypertension, cancer, mental illness) | *“Just like the name says, the diseases are not communicable. They cannot be passed from one person to the other. And most of* |
| NCDs |  | *them nowadays it's mostly about lifestyle."*<br>— FGD, HCW Nairobi. |
|  | Spiritual and cultural interpretations<br>(witchcraft, curses) | *"You will still find aspects of the beliefs, the traditional beliefs and stories about cancer patients that they have been bewitched... Mostly, it's associated with witchcraft."* — FGD, Youth Busia. |
|  | Stigma around cancer and mental illness; shame and misconceptions | *"Sometimes people fear associating with cancer patients, especially when they hear there's a smell or pus. They think it's contagious or shameful."* – FGD, Youth Busia |
|  | Cancer is feared and overwhelming | *"NCDs, some feel they are life-threatening... If someone hears that you have cancer, this person feels that you are dead the next day. So, this is one of the perceptions that is there."* – FGD, Youth Leaders, Nairobi. |
| 2. Socioeconomic determinants and lifestyle realities | Increasing NCDs among young people | *"We have the issues of arthritis and joint pain. Initially, we thought these were diseases that only affected older people, but* |
|  |  | *now we have people as young as 30 complaining of joint pains.” – FGD, HCW Nairobi.* |
|  | Recognition of modifiable risk factors | *“Eating healthy is good, but it’s expensive, especially for someone earning daily wages.” – FGD, Youth Busia.* |
|  | Socioeconomic constraints to healthy behaviors | *“...lack of resources to afford such kind of a meal because It requires a very high diet on vegetables...Most people, especially in Nairobi, are not able to get the correct vegetables because of the price” FGD HCW Nairobi* |
| 3. Health-Seeking Behaviors and System Barriers | Avoidance of health facilities due to cost, mistrust, distance, long queues | *“People try herbal remedies because they fear hospitals or can’t afford tests.” – CHP, Nairobi.* |
|  | CHPs as trusted educators and supporters | *“Every month, I can visit around 33 households or 35... If I come across someone who has a non-communicable disease, I advise.” – FGD, HCW Nairobi.* |
| 4. Perceived Opportunities and Limitations of AI in NCD Management | Lower AI familiarity among rural youth | *“We don't know who it is. I don't know, I haven't heard of it. There is this thing we used to love experiencing before WhatsApp, it was called Duta. Because duta knows everything...”*- FGD, Youth Busia |
|  | AI use for health information, reminders, symptom checking | *“... [AI] it will remind you, on things that you are not supposed to do and on things that you are to do, because it can reach a time whereby you can forget or you become tired so when you asking on AI, it can show you”* - FGD, CHP Busia |
|  | AI as support for reminders, referrals, tailored information | *“If AI could remind patients about meds and appointments, that would help us a lot. But it must be easy to use—even in the villages.”*<br>– FGD, HCW Nairobi. |
|  | AI as complementary to human delivered care rather than a replacement | *“It's good for information, but it cannot feel like a doctor does... sometimes you need that human touch.”* – FGD, Youth Nairobi. |
| 5. Ethical Concerns and Community-Centered | Concerns about privacy, data misuse, excessive data requests | *“Sometimes it asks for too much data—name, email, health history... you don't* |
| Implementation Pathways |  | *know who is seeing it.” – FGD, Youth Nairobi.* |
|  | Need for CHPs, youth, local leaders in AI education and rollout | *“We need our chiefs and CHPs to be trained too, so they can explain how these AI things work in our own language.” – FGD, Youth Busia.* |

### 1. Cultural and Biomedical Understandings of NCDs

Participants demonstrated awareness of NCDs as chronic, non-infectious conditions such as diabetes, hypertension, cancer, and mental illness. However, this biomedical understanding coexisted with spiritual and cultural interpretations especially in rural areas and NCDs are associated with stigma.

> *“Just like the name says, the diseases are not communicable. They cannot be passed from one person to the other. And most of them nowadays it’s mostly about lifestyle.”* — FGD, HCW Nairobi.
>
> *“You will still find aspects of the beliefs, the traditional beliefs and stories about cancer patients that they have been bewitched… Mostly, it’s associated with witchcraft.”* — FGD, Youth Busia.
>
> *“Sometimes people fear associating with cancer patients, especially when they hear there’s a smell or pus. They think it’s contagious or shameful.” – FGD, Youth Busia*.

Participants also described an increasing prevalence of NCDs among young people, contrasting with past assumptions that these diseases affected only older adults.

“*We have the issues of arthritis and joint pain. Initially, we thought these were diseases that only affected older people, but now we have people as young as 30 complaining of joint pains*.” – FGD, HCW Nairobi.

Cancer evoked particular fear, often described as a ‘death sentence’ regardless of stage. This fatalistic perception contributed to delayed care-seeking.

> *“NCDs, some feel they are life-threatening… If someone hears that you have a cancer, this person feels that you are dead the next day. So, this is one of the perceptions that is there.”* – FGD, Youth Leaders, Nairobi.

### 2. Socioeconomic determinants and Lifestyle Realities

While most participants recognized modifiable risk factors for NCDs, they consistently emphasized how poverty, food insecurity, and daily labor demands made it difficult to maintain these behavioral changes. Many participants only adopted healthier habits after receiving a diagnosis or experiencing a serious health scare.

> *“Financial, you’ve been told to use brown food, and you do not have money to buy them… sometimes like I don’t eat a brown ugali but because of the disease I’ll start to eat… so the first one is because of the finance, you don’t have money to use to buy.”* – FGD CHP Busia
>
> *“If someone is told to adopt an active lifestyle… this person is having a busy schedule, has to go to work and all that… by the time he or she comes from work, they are very tired. They are not able to do any physical activity.”* – FGD Youth Leader Nairobi

### 3. Health-System Navigation and Community Support

Despite general awareness of the need for early treatment, health seeking behavior was shaped by multiple barriers. These include cost, distance, mistrust of healthcare workers, long queues, and preference for herbal or traditional medicine.

> *“Other patients… tell you to tell the doctors to come and treat them where they are, because when they go there, there is queues. And what they don’t like is the queue in the facility that is a challenge.”*-FGD CHP Busia
>
> *“I don’t want to go to the hospital, because those girls will inject me. And they would even make me die earlier than I could… So, some have this notion… that he will die earlier when he goes there.” –* FGD CHP Busia
>
> *“People try herbal remedies because they fear hospitals or can’t afford tests.”* – FGD CHP, Nairobi.

Women were often reported to lack family support after diagnosis, increasing their vulnerability.

> *“I’ve seen lack of support from the other family members. For example, if it is one spouse, especially the women diagnosed with these NCDs, sometimes they really don’t get the support from their spouses.”* – FGD CHP Nairobi

CHPs emerged as trusted figures offering education and emotional support, especially in rural areas and serving as critical bridges between communities and formal health systems. However, participants noted their limited authority, training, and resources.

> *“We mostly do home visits, so when we do home visits and identified a problem, we do refer them. The referral does help them… when we visit them, they are always very happy also it helps us to know a lot of things.”* – FGD CHP Busia
>
> *“What make it easier is being very close to them… you make stories until he forgets that this person is so close to me… so that he can call me… and he can go to get treatments.”* – FGD CHP Busia

### 4. Perceived Opportunities and Limitations of AI in NCD Management

Awareness and use of AI tools varied across urban and rural areas. Youth and some healthcare providers in Nairobi described using platforms such as ChatGPT, symptom checkers, and health apps for reminders or information. In contrast, participants in Busia, which is more rural, had limited exposure to AI, reflecting broader gaps in digital literacy and access between rural and urban areas. Despite this gap, youth and CHPs in both urban and rural areas identified potential applications of AI for; disease diagnosis and detection; health monitoring and fitness tracking; health information and symptom checking health; data storage & management; nutrition & lifestyle management applications; and healthcare delivery and service automation.

Simplicity, cultural relevance, and ease of use were emphasized as critical for adoption and impact.

> *“AI can be made to work in different circumstances… you won’t need the internet to access information about AI. Let it be like in your phone… So, AI should be like that at least it would help a lot of people.”-* FGD Youth Busia

### 5. Future Recommendations: Addressing Ethical Concerns and Community-Centered Implementation Pathways

Concerns about accuracy, privacy, data collection, and excluding low-literacy or low-connectivity populations were raised. Consent and transparency were seen as essential for trust. Participants emphasized the significance of consent, data protection, and equitable design in AI implementation.

> *“… if you use AI, yes, it is cheaper than going directly to the doctor, but it will still give you wrong information. Treating something that you aren’t sure. So I don’t see that it is okay”-* FGD Youth, Nairobi
>
> *“Privacy. To me, that’s the challenge when using AI because sometimes there are leakages in terms of database… we have very confidential information that can be leaked… there are leakages in AI.”* – FGD Youth Nairobi
>
> *“So, AI can be prone to hackers… it is not safe… some sensitive information can get lost.”* – FGD Youth Nairobi
>
> “*Sometimes it asks for too much data—name, email, health history… you don’t know who is seeing it.”* – FGD, Youth Nairobi.

Access issues: low literacy, language barriers, and technological limitations can exclude some populations

*“Most of the people in my community, they don’t know how to read and write, they don’t know how to use these things, these smartphones… most of the people maybe they know their mother tongue. So how will they type?”* – FGD Youth Leaders Busia

### 6. Community-engaged approaches and policy mobilization recommended

Participants called for inclusive, community-driven approaches to education and policy, highlighting the need to engage youth, CHPs, and local leaders in the design and rollout of AI health tools. Broader community education on both NCDs and AI was seen as essential, with trusted messengers playing a central role in ensuring understanding and uptake. Participants also urged policymakers to invest in digital infrastructure, expand digital literacy, and ensure that marginalized populations are not left behind in health innovation efforts.

“*We need our chiefs and CHPs to be trained too, so they can explain how these AI things work in our own language.”* – FGD, Youth Busia.

> *“It would be better if we could have something I would call total youth involvement in that if project design up to the implementation stage, its involvement also decreases so we actually own it as the young people… So, if we fighting a disease and we are involved on the last minute, why don’t we be at the forefront so that we can start fighting it early?”* – FGD Youth Leaders Busia

## Discussion

This was a qualitative assessment of a youth population in Kenya with engagement and attitudes towards NCDs and AI for health previously unassessed. Overall, we found an encouraging level of awareness of NCDs among youth in this population. However, this is hampered by deep-rooted stigma, spiritual beliefs, cultural perceptions, and economic challenges that govern behavior and delay access to care. There was a varied awareness of emerging AI technology with participants in urban areas exhibiting greater awareness of this technology and ways it could be applied to the prevention and management of NCDs.

Specific to NCDs, it was evident that spiritual and cultural interpretations, such as beliefs in witchcraft, curses, and evil spirits, continue to shape how NCDs are understood and managed, especially in rural areas. These beliefs often delay care and foster stigma, particularly around cancer and mental illness. Stigma manifests as social isolation, shame, and fear, deterring individuals from seeking timely care and contributing to late diagnoses and poor outcomes (11–15).

The adoption of lifestyle changes associated with prevention of NCDs is dismal. Despite increased awareness, participants reported that economic challenges, poverty, food insecurity, and daily labor demands make it difficult to adopt and sustain healthy behaviors. Many only changed their lifestyle after a diagnosis or health scare. Barriers to care included cost of care, distance to facilities, mistrust, and long queues. These issues are not uncommon in the region; additional studies in Kenya have shown similar patterns of high healthcare costs and access barriers. Similarly, in nearby Uganda, patients endorse identical concerns about unaffordable medical bills, transportation costs, long waiting times, and the absence of healthcare workers (16). Some solutions include enabling self-care and decentralized health care clinics, with successful models demonstrating feasibility of community-based integrated care for NCDs through differentiated service delivery approaches and task-sharing with non-physician clinicians (17). Similarly, in our study, AI and digital tools were viewed as promising, across all categories of participants, for their potential to support self-care through applications that promote medication adherence, symptom awareness, and health education, representing a novel and encouraging perspective in contemporary healthcare contexts (18)

Herbal and traditional medicine use remains pervasive, augmented by barriers to clinical facilities and beliefs about intrinsic efficacy of traditional remedies. However, this can cause delays in timely treatment, result in delayed diagnosis and contribute to poor outcomes for conditions requiring immediate biomedical intervention (19,20).

Participants identified CHPs as essential connectors between health systems and communities with a critical role in facilitating initial NCD care, as they have widespread access to a large number of community members; they provide health education, emotional support, and key initial screening and education. However, their successes in NCDs prevention and management is often constrained by a lack of authority, insufficient resources, and inadequate training(21). Further structural barriers on the part of the patient such as high costs, geographical distance, long queues, and mistrust in the healthcare system lead to failed referrals). Since CHPs are trusted and effective for initial engagement, policymakers should continue to ensure that CHPs have appropriate training, infrastructure, and resources to support and refer for chronic disease care.

. Although participants were optimistic about the potential of AI-powered apps for NCDs management through such as reminders and symptom checking, and the adoption of WhatsApp-based engagement platforms, digital inequality and mistrust in data use pose substantial challenges. Aspects of digital inequality that were highlighted include limited access to smartphones, high internet costs, and low digital literacy, which remain substantial barriers, especially in rural areas). This reflects previous findings showing that while digital health interventions are generally feasible and acceptable among youth in LMICs, persistent digital divides caused by cost, infrastructure, and digital literacy must be addressed to ensure equitable access (22–24). Importantly, participants emphasized that successful implementation of AI requires simple, localized, and culturally attuned tools that complement, but do not necessarily replace human care. Furthermore, in existing literature, digital health chatbots need to be adapted for the given context – including language options and familiar content; the AGILE chatbot for gender-based violence services in Kenya is one example that demonstrates the importance of user preferences and cultural adaptation in digital health tool development by adopting a human-centered approach in designing an effective chatbot with as many human features as possible to optimize utilization (25). In summary, AI-based tools provide a promising solution for health among Kenyan youth but will need to be adapted to the local context accounting for barriers in access. Advanced digital education can be employed to target youth in these contexts, including early in schools and at universities, to increase capacity for engagement (26).

## Limitations

This study is limited by its sample size and geographic focus on only two counties, which may not represent national trends. Focus groups may also be subject to social desirability bias as there was no verification of actual AI use. However, the diversity of participant backgrounds offered rich contextual insights, and we ensured purposive selection for reorientation across key domains such as gender, residence, and age.

## Conclusion

This study highlights the ongoing challenges and emerging opportunities in addressing NCDs in Kenya. While public awareness of these conditions is increasing, many individuals still face barriers such as cultural stigma, economic hardship, and systemic healthcare limitations. Each of these factors impedes early detection and effective management of NCDs.

AI offers a promising opportunity to improve current health services, particularly in underserved areas and among youth especially in addressing modifiable risks of NCDs. This research shows that there is already ongoing spontaneous adoption of AI technologies such as Large Language Models (LLMs) for nutritional counselling and the use of wearables to track physical activity. Incorporating new technologies in healthcare must consider the realities of the communities they serve, ensuring compatibility with local languages and cultural norms. It should also optimize access to care. Integrating AI into trusted, community-based platforms through both individual direct to consumer applications and through CHP toolkits improves care delivery, strengthens health education, and enhances treatment adherence.

Future research should expand culturally relevant health education and address barriers related to digital and health literacy. Policy frameworks must prioritize data privacy, ethical safeguards, and equitable access to digital health resources. By integrating digital innovation into established community health structures, Kenya can cultivate a more inclusive, resilient, and sustainable approach to preventing and managing NCDs.

## Data Availability

All data produced in the present study are available upon reasonable request to the authors.

## Acknowledgments

Report any acknowledgments here/None.

## Footnote

Reporting Checklist: The authors have completed the SRQR reporting checklist (27).

## Funding

This research was funded by Gates Catalyzing Equitable AI Use Grant

## Conflicts of Interest

None

## Ethical Statement

## Notes

### Competing Interest Statement

The authors have declared no competing interest.

### Author Declarations

The study received ethical approval from the African Medical and Research Foundation (AMREF) Ethics and Scientific Review Committee (Ref: ESRC P1518/2023), National Commission for Science, Technology and Innovation (NACOSTI/P/24/31792), and County Health Offices in Nairobi and Busia.

